# Individual costs and societal benefits of interventions during the COVID-19 pandemic

**DOI:** 10.1101/2023.02.08.23285651

**Authors:** Arne Traulsen, Simon A. Levin, Chadi M. Saad-Roy

## Abstract

Individual and societal reactions to an ongoing pandemic can lead to social dilemmas: In some cases, each individual is tempted to not follow an intervention, but for the whole society it would be best if they did. Now that in most countries the extent of regulations to reduce SARS-CoV-2 transmission is very small, interventions are driven by individual decision-making. Assuming that individuals act in their best own interest, we propose a framework in which this situation can be quantified, depending on the protection the intervention provides to a user and to others, the risk of getting infected, and the costs of the intervention. We discuss when a tension between individual and societal benefits arises and which parameter comparisons are important to distinguish between different regimes of intervention use.

## 1. Introduction

The SARS-CoV-2 pandemic is an ongoing public health emergency that has led to significant morbidity and mortality (*e.g*. see [1]). Since the beginning of the pandemic, individuals across the world have implemented various measures to prevent spread. These approaches began with nonpharmaceutical interventions (NPIs), such as mask-wearing, school closures, and widespread lockdowns. The advent of effective vaccines (*e.g*. [2, 3]), *especially against severe disease (e.g*. see [4]), generated a transition away from NPIs toward vaccination as a pharmaceutical intervention (PI). However, the emergence of novel SARS-CoV-2 variants capable of immune escape (*e.g*. [5]) illustrates the importance of continued adjustments to any decision (whether personal or societal) aimed at decreasing transmission and preventing rapid exponential growth.

Many of the NPIs and PIs used to prevent transmission have individual and societal impact. For example, since society would benefit from decelerated epidemic spread, individuals could decide to reduce their social contacts at personal costs. Many scientists have argued that this leads to social dilemmas [6–12], where the individual optimum is in conflict with the societal optimum. A social dilemma is a situation in which “decisions that make sense to each individual can aggregate into outcomes in which everyone suffers” [13]. For the interaction between two players, this can be illustrated by a payoff matrix

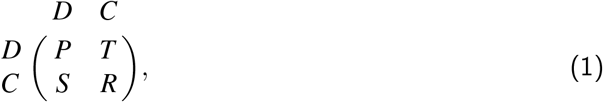

where e.g. *T* is the payoff of a player defecting (*D*) against a player that cooperates (*C*). For *R* > *P*, players prefer mutual cooperation over mutual defection, but if *T* > *R* either player is tempted to defect (“greed”), whereas for *P* > *S* a player would prefer to defect against another defector (“fear”) [13]. For *T* > *R* > *P* > *S*, we have a Prisoner’s Dilemma. For *T* > *R* > *S* > *P*, we have a Snowdrift [14] or Chicken game [13]. For *R* > *T* > *P* > *S*, we have a Stag Hunt Game [13, 15].

Before and during the COVID-19 pandemic, such social dilemmas have been discussed e.g. in the context of social distancing [16, 17], mask usage [7, 8, 18, 19], and vaccination [20–22]. Here, we argue that whenever the intervention leads to higher benefits for others than for the individual conforming to them, there is a generic possibility of a social dilemma – but depending on the individual costs for the intervention and the current state of the pandemic, there may also be no social dilemma at all. We argue that when individuals do not take the future development into account, a social dilemma only appears for intermediate disease loads in the population. In times of very high or very low risk, the individual optimum is fully in line with the social optimum. In this case, the enforcement of interventions is less challenging, as the imposed rules are consistent with the individual decisions of individuals. However, for intermediate disease loads, individuals are tempted to deviate from the social optimum and such a regulation can lead to conflicts – and measures that put individual protection into the focus are more likely to become popular.

We begin by introducing a simple framework that examines the benefits of interventions in conjunction with individual and societal costs. We analyze the model, giving intuition for potential outcomes at the individual and societal levels, and illustrate the use of our framework with three potential interventions. We then extend our framework to include a situation where individuals may choose among different interventions, each with their distinct cost and benefit.

## 2. Framework formulation

We first examine a payoff matrix in which a focal individual can decide to follow a given intervention. This focal individual is interacting with other individuals facing exactly the same choice. We assume that the probability that somebody they interact with is infectious with COVID-19 is Ξ. In our model, Ξ will be constant, which can be assumed for short-run interventions. In the long run, the course of the pandemic is intertwined with the decisions of individuals to follow interventions [23–29]. A more realistic model would have to take into account this intertwinement, but also the mechanisms of infection and the associated time scales. We assume that the cost of getting infected is *ξ*. This number may be different for each individual, as it depends on the health status of that person and also on the personal living situation (do they live with others that are at a higher risk?) and short term plans (*e.g*. such as important personal events). However, we assume for simplicity that both interacting individuals have the same *ξ*. In our model, only the product Λ = Ξ *ξ*, which captures the probability to get infected and the risk associated to it, matters.

In addition, we assume that a fixed cost *γ*_*I*_ is associated to the intervention, for example the personal costs of decreasing social contacts or the perceived personal costs of vaccination. Note that in general, *γ*_*I*_ can depend on how many individuals are observing the intervention, for example in the case of social ostracism of mask wearers or of non-wearers. This leads to the following payoff matrix for each interaction, where individuals can follow the intervention (strategy *I*) or not (strategy *N*),

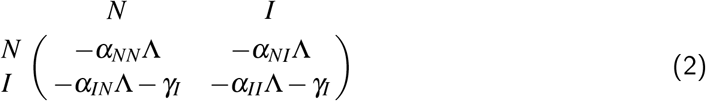

The entries in the matrix are the payoffs that the focal player obtains by interacting with the other player. For example, if they choose *N*, their payoff is −*α*_*NN*_Λ if the other player does not follow the intervention and −*α*_*NI*_Λ if the other player follows it. In this matrix, *α*_*NN*_ measures the risk of infection from an interaction with an infectious individual if neither is following the intervention. We assume that interventions come with individual benefits, reducing the probability *α*_*NI*_ that the focal individual is infected when it follows the intervention, *α*_*NI*_ < *α*_*NN*_. In addition, interventions also come with societal benefits, reducing the probability that others are infected in this case, *i.e. α*_*NI*_ < *α*_*NN*_. Finally, in the absence of costs, when all individuals adhere to the intervention the probability to get infected will be lower than in a situation where nobody follows an intervention, *i.e. α*_*II*_ < *α*_*NN*_. Here, we focus on the case where adhering to an intervention benefits the user less than the interaction partner, *α*_*NI*_ < *α*_*IN*_ and where the optimum occurs when both adhere to the intervention, which leads to

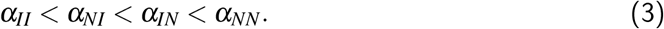

Note that the ranking (3) does not yet determine whether there is a social dilemma or not – this is established by this ranking, whether the condition *α*_*NI*_ − *α*_*II*_ < *α*_*NN*_ − *α*_*IN*_ is fulfilled, the general risk in the population, and the cost of the intervention.

First, we examine the outcomes qualitatively, thinking of an individual that interacts with many others in a population. If there is either high incidence, or a high risk from the disease compared to the cost of the intervention (*i.e*. Λ ≫ *γ*_*I*_), adhering to the intervention is a dominant strategy: The payoffs in the first line of the payoff matrix (2) are always smaller than the payoffs in the second line due to −*α*_*NN*_ < −*α*_*IN*_ and −*α*_*NI*_ < −*α*_*II*_. In this case, there is no conflict between the personal and the social optimum.

If there is either low incidence or a low risk from the disease compared to the cost of the intervention (*i.e*. Λ ≪ *γ*_*I*_), not adhering to the intervention is a dominant strategy. This is because the payoffs in the first line of the payoff matrix (2) are driven by the costs and thus always larger than the payoffs in the second line for Λ ≪ *γ*_*I*_ – also in this case, there is no conflict between the personal and the societal optimum. Only for intermediate Λ can a social dilemma arise.

However, if the pandemic coevolves with the intervention, the situation may get worse if people do not follow interventions. In this case, individuals who optimize their payoff in the present without taking into account the future may be worse off later on. This would require a more complex model in which the game can change with time [30]. Here, we assume instead that our interventions occur on a time scale that is short enough to abstract from such complications.

We now assume that the interaction with many individuals arises many such games, such that payoffs depend on the fraction of other players following the intervention. We also assume that the payoff from individual interactions are additive, which is in our case reasonable as long as the risk of an infection is not too high – otherwise, an individual would care less about additional interactions if their personal risk after a few interactions is already very high. We also assume that individuals are only interested in their individual payoff. In most cases, we cannot assume that players will maximize their payoff by analyzing the game in detail. Instead, we assume that players initially use different strategies, leading to payoffs that depend on the composition of the population. Then, players imitate predominantly those that have high payoffs [31–33], eventually leading to either a homogeneous population where everyone follows the intervention, a homogeneous population where nobody follows the intervention, or a mixed population of followers and non-followers. A popular choice for this is the replicator dynamics, which we also use (see Appendix A).

Let us now analyze the game more quantitatively. We analyze the game starting from a situation with the highest risk and ask when the payoff structure changes qualitatively. For simplicity, we focus on the case where the additional protection that is obtained by switching to the intervention when an interaction partner already follows, *α*_*NI*_ − *α*_*II*_, is smaller than the additional protection that is obtained by following the intervention when the interaction partner does not follow, *α*_*NN*_ − *α*_*IN*_ (see Appendix B for the opposite case). The imitation process leads to the following outcomes:

1. When 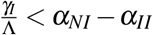, the situation is so risky that adhering to the intervention is optimal for all individuals, resembling a “Harmony game” in game theory.
2. For 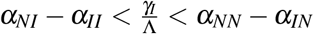, we have a Snowdrift game: If everybody is adhering to the intervention, it is fine for an individual to stop. If nobody is adhering to the intervention, an individual should start. In a large population, this would lead to a stable fraction 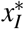 of individuals adhering to the intervention

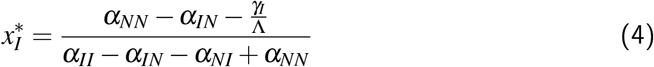

In this situation, universal adherence is the social optimum, but individual decision making leads to less adherence.
3. For 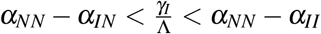, we have a Prisoner’s dilemma – adhering to the intervention is dominated by not adhering, but the social optimum is still that everyone adheres to the intervention.
4. For 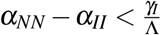, adhering to the intervention is entirely dominated by not adhering – either because the incidence Ξ is low or because the cost *ξ* is low.

Note that when the ranking of our *α* parameters is different, some of these regimes can be absent. For example, the Prisoner’s Dilemma region would only vanish if the protection of somebody following the intervention is better than the mutual protection in a situation where both follow the intervention – an unlikely scenario.

## 3. Examples

### 3.1 Reducing social contacts

Social distancing is an important intervention, which in many cases fulfils the ranking in (3): The risk becomes lowest when everybody reduces contacts and is highest if everybody maintains many contacts.

Let us look at a concrete example for the reduction of social contacts: Assuming that the cost of an infection Λ = 1 and that the cost of reducing social contacts is of a similar magnitude, *γ*_*I*_ = 0.9. In addition, we set the *α* parameters to *α*_*NN*_ = 1.0, *α*_*NI*_ = 0.9, *α*_*IN*_ = 0.2, and *α*_*II*_ = 0.01. We assume that these parameters capture the effect of the assortment that naturally arises: Individual who do not distance from others would naturally have more contacts with each other and thus be more likely to be infected, increasing the risk of those interacting with them. This leads to a payoff matrix

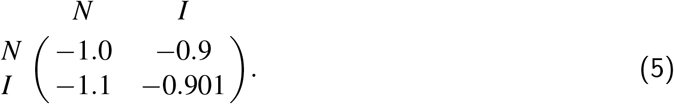

As the payoffs in the first line are always higher than the payoffs in the second line, not reducing social contact is a dominant strategy. However, this leads to a situation that is socially suboptimal, as the payoff in a homogeneous populations where nobody reduces social contacts (−1.0) is lower than the payoff in a population where everyone would reduce social contacts (−0.901).

In most realistic cases, however, individual will have different risk assessment (e.g. they may be prone to more severe disease due to pre-existing conditions) and they will experience different costs of interventions (reducing social contacts may be easy for some and very difficult for others). Let us assume that two individuals interact and that the cost of reducing social contacts is *γ*_1_ = 0.9 for the first one and *γ*_2_ = 0.5 for the second one. All remaining parameters are as in the numerical example above. This leads to the payoffs

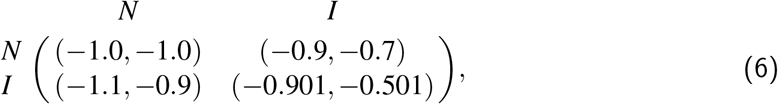

where the first player is the row player (and obtains the first payoff) and the second player is the column player (and obtains the second payoff). Now the situation where both do not follow the intervention, (*N, N*) with payoffs (− 1.0, −1.0), is no longer an equilibrium. Instead, the second player will start following the intervention, as it increases their payoff, leading to (*N, I*) with payoffs (− 0.9, −0.7).- In this asymmetric situation, the second player with the lower cost will follow the intervention and reduce their contacts, while the first one maintains their contacts. The first player benefits from the second player following the intervention, but has no incentive to follow herself. In a more realistic scenario where also the other parameters are different (e.g. the *α* parameters could be different for the two players), such asymmetry would be a generic, but apparently unfair outcome: Those who assess the risk as higher and those who have lower costs to reduce their contacts are more likely to follow interventions, improving the situation for both at an individual cost.

### 3.2 Vaccination

The benefits of vaccination can come in different forms: They can reduce transmissibility to others and they can reduce susceptibility for the vaccinated individual [34]. They can also just reduce the probability of severe disease and not have a direct impact on the course of the pandemic. The example of vaccination is most likely to violate our assumption of constant overall risk Ξ, as the benefits of vaccination occur on a longer time scale compared to e.g. the usage of masks.

Let us start by thinking of a hypothetical vaccine that only reduces the transmissibility to others. This means that individuals benefit from the vaccination of others, but not from the vaccination of themselves. It leads to the ranking

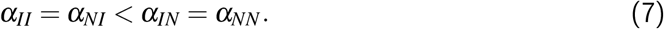

In this case, for low costs of vaccination or high risks of infection, 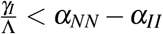, we have a Prisoner’s Dilemma – individual decisions would lead to no vaccinations, but for every individual it would be beneficial if others are vaccinated. For high costs or low risks, 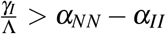, vaccination costs would outweigh the benefits and individuals would not get vaccinated – such a case is particularly likely if there is a high perceived costs of vaccination (the real costs of vaccination tend to be very low [35]). Thus, vaccine usage would expected to be minimal if it only reduces transmissibility to others, despite its societal benefits in risky situations.

Next, let us turn to a hypothetical vaccine that only reduces the susceptibility for the person vaccinated. This means that individuals benefit from the vaccination of themselves, but not from the vaccination of others. It leads to the ranking

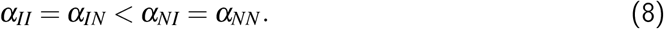

In this case, for low costs or high risks, 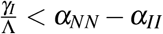, everybody would get vaccinated. For high costs or low risks, 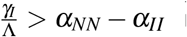, nobody would get vaccinated, as the costs outweigh the benefits. There is no social dilemma in this case, as the benefits of vaccination would be purely individual.

A true vaccine will reduce both transmissibility and susceptibility, such that we would expect

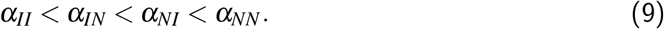

In this case, we recover all the four cases discussed above for the general intervention scheme. A hypothetical vaccine that only reduces the probability of severe disease would have no direct impact on the dynamics of an epidemic, as the same number of cases and transmissions would be observed – but the cost of getting infected, *ξ*, would decrease for the vaccinated, leading in turn to a different value for the overall risk Λ, potentially changing the behavior. Since they would at first only confer individual benefits, the uptake of such a vaccine would depend solely on individual assessments.

If a durable transmission-blocking vaccine is deployed widely, community immunity would decrease local infections and potentially lead to local elimination (see, [24, 25, 36–38]). In turn, individuals would perceive their individual risk of infection if they remain unvaccinated to be low, and thus the ratio of individual risk to cost would change. With waning vaccinal immunity, infection levels could rise again, and thus alter decision-making.

In the case of vaccination, one would expect that the real risk of an infection differs widely between individuals due to their age and health status. However, in a situation where the cost of vaccination is very low, one would still expect that everyone would get vaccinated. But on the other hand, social norms and group dynamics as well as spread of misleading information on vaccination costs can lead to to situations where vaccine uptake becomes very heterogeneous [35].

### 3.3 Mask usage

Recent studies have underlined the importance of extending epidemic models to include the dynamics that surround the social norms of mask-wearing [23, 28]. We can use our simple framework to gain intuition into the choices individuals make for different kinds of masks.

The simple cloth masks that were initially promoted in most countries during the pandemic mostly served to protect others – so one would expect that they naturally fulfill the ranking in (3). While this has been discussed widely in science and also on social media, it is challenging to quantify this, as controlled infection experiments are not feasible. However, as a proxy one can take aerosol measurements exhaled from people without masks or with different kind of masks.

In a situation where everyone wears a surgical mask, should people switch to masks that offer better protection, such as KN95 or FFP2? Bagheri et al. [39] have performed a study measuring aerosols in such a context. Denoting the simple surgical mask by *M* and the higher quality mask by *F*, the relevant parameters they measured for our context are *α*_*FF*_ = 0.0014, *α*_*MF*_ = 0.0097, *α*_*FM*_ = 0.015, and *α*_*MM*_ = 0.104 (cf. Fig. 6, columns *FF, FS, SF*, and *SS* in [39]). Note that the ranking *α*_*FF*_ < *α*_*MF*_ < *α*_*FM*_ < *α*_*MM*_ is satisfied here. Thus, choosing between the two kinds of masks is an example for the situation depicted in Fig. 1.

**Figure 1:**
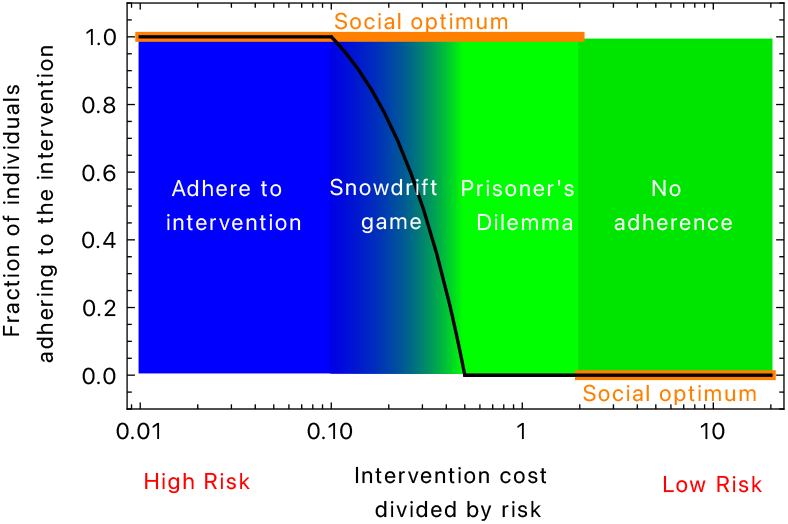
Fraction of individuals that are expected to adhere to an intervention. The black line indicates the equilibrium fraction individuals adhering to an intervention, the orange line shows the social optimum. Whenever these two lines deviate from each other, interventions to enforce a social optimum would have an influence on individual behavior. If the social optimum is identical to the equilibrium fraction, interventions do not have to be enforced (parameters: *α*_*NN*_ = 3.0, *α*_*IN*_ = 2.5, *α*_*NI*_ = 1.1, *α*_*II*_ = 1.0).

A very different situation arises for masks that protect *only* the user, e.g. about KN95 masks with vents, which allow free circulation to the outside, but filter the air inhaled by the user. We denote using these masks by *V* and first look at the situation where individuals choose between these masks and no masks. For these masks, we have *α*_*VV*_ = *α*_*VN*_ < *α*_*NV*_ = *α*_*NN*_. Thus, only two situations are possible:

1. For 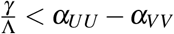, it is the best option for everyone to stick to *V* masks.
2. For 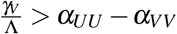, nobody would wear a mask, as the cost of wearing a mask is higher than the benefit from it.

Finally, let us discuss the case where individuals can choose between two different interventions. We focus on choosing between two different kinds of masks. Consider a case where there are no mask mandates, so individuals can freely choose between wearing no mask (*U*), simple surgical masks (*M*) and higher quality masks (*F*) that tend to cause the highest costs. First, we need to quantify our *α* parameters. There are several sources that give such numbers, but often their origin is unclear and a close inspection reveals inconsistencies (for example, it is unlikely that any type of mask gives equal protection, regardless whether it is used by the infected or the susceptible individual). Thus, we again use the study by Bagheri et al. [39] and estimate the protection levels of individuals not using any mask, see Table 1.

**Table 1:** Parameters for the protection levels of two different masks and unmasked individuals based on the study by Bagheri et al. [39]. The five parameters that are not given in that study are estimated according to the following assumptions: ^a^No mask is twice as risky as an *M* mask for both interactions, leading to a risk increase by a factor of 4 comparing *α*_*UU*_ to *α*_*MM*_. ^b^The risk reduces more if an infected individuals wears an *M* mask (*UM*) compared to an uninfected individual trying to protect herself with an *M* mask (*MU*), *α*_*UU*_ > *α*_*MU*_ > *α*_*UM*_ > *α*_*MM*_. ^c^ The risk reduces more if an infected individuals wears an *F* mask (*UF*) compared to an uninfected individual trying to protect herself with an *F* mask (*FU*), *α*_*UU*_ > *α*_*FU*_ > *α*_*UF*_ > *α*_*FF*_. In addition, we assume wearing no mask interacting with an infected *F* mask wearer leads to higher risk than using an *F* mask when the infected one uses an *M* mask, *α*_*UF*_ > *α*_*FM*_. ^d^ Following the assumption we made for *α*_*UU*_, my risk doubles if the infected one removes mask. ^e^ If I wear an *F* mask, my risk increases slightly when the infected one switches from *M* to *U* (*α*_*FU*_ > *α*_*FM*_).

| Parameter | Source |
| --- | --- |
| $\alpha_{UU} = 0.40$ | estimated<sup>a</sup> |
| $\alpha_{UM} = 0.15$ | estimated<sup>b</sup> |
| $\alpha_{UF} = 0.018$ | estimated<sup>c</sup> |
| $\alpha_{MU} = 0.20$ | estimated<sup>d</sup> |
| $\alpha_{MM} = 0.104$ | Fig. 6, column SS in [39] |
| $\alpha_{MF} = 0.0097$ | Fig. 6, column FS in [39] |
| $\alpha_{FU} = 0.02$ | estimated<sup>e</sup> |
| $\alpha_{FM} = 0.015$ | Fig. 6, column SF in [39] |
| $\alpha_{FF} = 0.0014$ | Fig. 6, column FF in [39] |

With this, we can analyse the situation as a 3 × 3 game,

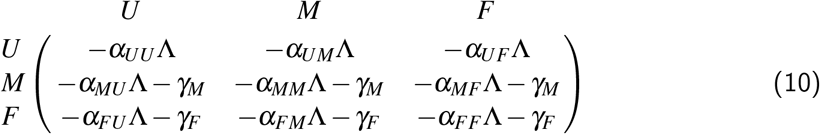

Here both masks have different benefits and costs. For *γ*_*F*_ ≫ *γ*_*M*_, the *F* masks are irrelevant and the analysis reduces to the case of *U* vs *M* masks. Similarly, for *γ*_*F*_ ≪ *γ*_*M*_, the *M* masks would be irrelevant and the analysis reduces to the case of *U* vs *F* masks. A full overview of the expected mask usage in such a case is given in Fig. 2.

**Figure 2:**
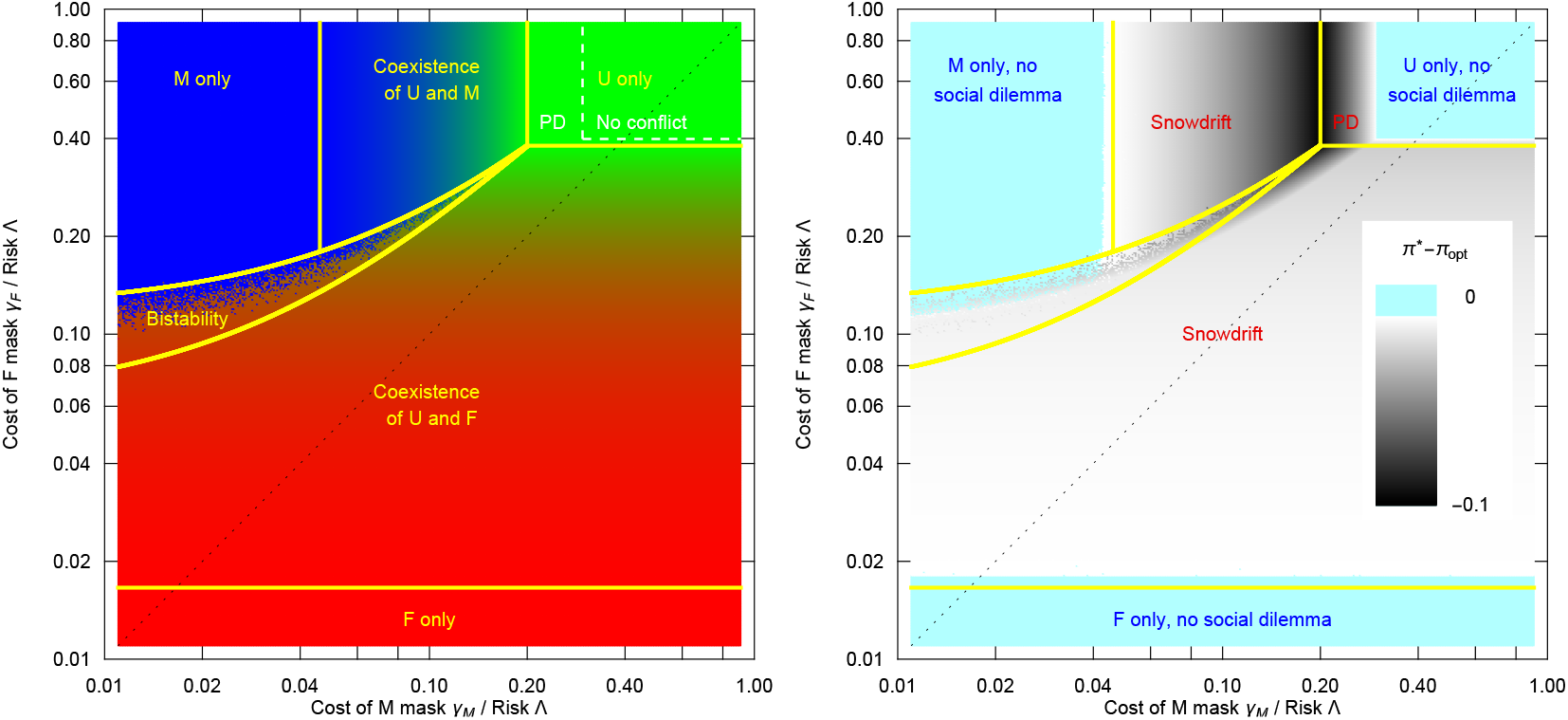
Choosing between lower quality masks (*M*) and high quality masks (*F*). Left: Depending on the costs of using either the lower quality masks (*γ*_*M*_) or higher quality masks (*γ*_*F*_) and the risk situation determined by Λ, we have a number of different scenarios in terms of mask usage: For *γ*_*M*_ > *γ*_*F*_ (lower right), the lower quality masks play no role and we recover a situation where for a wide range of parameters, there is a mix of people using masks and people not using masks. When the risk is reduced (moving from bottom to top), this turns into a Prisoner’s Dilemma like situation and for very low risks, not wearing a mask is dominant. For *γ*_*M*_ < *γ*_*F*_ (upper left), the situation is more interesting: For very large *γ*_*F*_, all scenarios from universal mask wearing to no mask wearing are recovered, cf. Fig. 1. But for *γ*_*M*_ ≈ *γ*_*F*_, wearing a more costly mask can be beneficial in situations where simple masks would be worn only at a low frequency. Consequently, there is a large region of coexistence between *U* and *F*, with the fractions of the two types given by the equivalent of Eq. 13 (replacing the parameters of *M* masks by those of *F* masks). In addition, there is a region of bistability, where two solutions are possible: Either the use of *M* masks by everyone or a coexistence between *F* and *U*. Right: The payoff difference between the situation emerging in our model, *π*\*, and the social optimum, *π*_opt_. Dark areas depict a large difference, i.e. a strong social dilemma. The dilemma is particularly strong when high quality masks come with high costs. However, a Prisoner’s dilemma (PD) arises in a small region of this space only, the typically dilemma represents a Snowdrift game (parameters as in Table 1, see Appendix A for details on this figure).

In particular for intermediate costs, there are additional interesting cases discussed in Appendix C. For example, there is a region of the cost parameter space in which there is a bistability: The whole population would either use *M* masks only or there would be a coexistence between the use of *F* masks and no masks *U* – and the initial state would determine which solution is reached. For the parameter set used, the two types of masks do not coexist, see Fig. 2.

## 4. Discussion and conclusion

The ongoing SARS-CoV-2 pandemic has illustrated the importance of individual behavior in pandemic response. Here, we have formulated and analyzed a basic game-theoretic framework that examines the individual costs and societal benefits of interventions. We have illustrated the use of our framework through three examples: social distancing, mask-wearing, and vaccination. Each of these highlight how our model can be used as a guide to understand individual choices through the pandemic. These examples also underline the importance of developing strategies with characteristics (parameter regimes) such that individual decisions are aligned with positive population outcomes.

Our framework makes a number of assumptions, which should be examined in future work. For example, we assume a constant level of infections. In reality, this is dynamic, and epidemic trajectories could contribute to individual decision making regarding multiple interventions. In turn, these decisions could themselves impact epidemiological dynamics. Studying these implications is an important area of current and future research [23–29]. We have also assumed that individuals are acting only in their own short term self interest – but in reality, some may also act in the interest of their community or in their own future interests.

We have focused on three different examples of interventions: social contacts, vaccination, and masks, and discussed these individually. However, individuals may choose to adhere to a combination of these interventions, each with different perceived costs. While analyzing the game-theoretic outcomes in this case is substantially more complicated, the advent of more data on these interventions may eventually enable such an endeavor.

Additionally, we have used a unifying framework to examine both PIs, such as vaccination, and NPIs. In reality, NPIs and PIs can be very different – for example, vaccination has an effect that lasts at least months, whereas mask usage can be changed very quickly. In addition, NPIs and PIs may have very different perceptions at the population level, which can also affect individual behaviour. Consequently, there can be feedback between perceived individual risks and population-level benefits and thus individual behaviour [22]. Future work should extend our framework to examine these complexities.

Furthermore, we have omitted many complexities involved with vaccination. For example, multiple vaccines were initially deployed as two-dose vaccines, and potential changes in individual decisions regarding the second dose were discussed [40, 41]. The advent of subsequent booster doses (with varying uptake levels) further reveals the importance of individual decisions on a per-dose basis. Thus, extending our model to include multiple doses would allow us to capture the potential change in social dilemma that emerges. In turn, this would be very useful to understand individual decision-making in the face of multi-dose vaccines.

Maybe most importantly, individuals differ in their individual risk assessment and in their risk preferences. This implies that some individuals perceive the situation as one where it would be in the individual interest of everybody to follow an intervention, while others perceive it as a social dilemma or even a situation where following interventions is no longer necessary and they are no longer willing to follow an intervention. Such a heterogeneous risk assessment makes a game-theoretical analysis much more challenging, but it is important to consider this case: As discussed in the example of social distancing, different individual assessments can lead to outcomes that are perceived as unfair, as those who perceive higher risk or have lower costs following an intervention will follow them, but not those who perceive lower risk or have higher costs following an intervention. Thus, individual heterogeneity is one potential source of social tensions arising from a pandemic, especially when large groups of the population emerge that have fundamental disagreements about risk assessment [42]. In particular, for vaccination, risk perception may be a more important driver of decision-making, and hesitancy towards vaccination may spread as a contagion [35]. Finally, beyond individual assessment and preferences, there could also be heterogeneities in risk, *e.g*. there could be a vulnerable group within a population. This could be partly addressed by extending the model to different demographic or social groups, where e.g. the elderly are at a higher risk or medical personal is exposed at a higher rate than others. Such a situation could lead to stronger social tension, as the societal benefits of adherence to an intervention would increase for part of the population. Future work should examine these implications in detail.

Relatedly, we have also assumed that the cost of intervention is the same across the population. However, heterogeneities, such as in age or space, could impact both infection risk and cost of intervention. Extending our model to investigate the impact of underlying heterogeneities on individual behaviour would be important.

Overall, our findings highlight the importance of characterizing (and parameterizing) the costs and benefits of each intervention as they are proposed. In the absence of government-imposed regulations, our framework illustrates the impact that these costs and benefits can have on individual-level decision making. In turn, these decisions will be central to determine the future course of the pandemic. Thus, the intuition gained by our simple framework can guide policy-makers as they decide whether individuals would adhere to a proposed intervention. In addition, it helps to grasp some of the roots of conflicts about following interventions or not when risk preferences and cost assessments are heterogeneous.

## Data Availability

All data produced in the present work are contained in the manuscript

## Acknowledgements

AT thanks Corina Tarnita at the EEB department at Princeton University hosting him on a sabbatical, where this work was initiated. We thank Bryan Grenfell and Christian Hilbe for stimulating discussions and comments on an earlier draft. AT acknowledges generous core funding by the Max Planck Society. SAL acknowledges funding by the NSF (grants CCF-2142997, CNS-2041952, and CCF1917819). CMSR acknowledges funding from the Natural Sciences and Engineering Research Council of Canada via a Postgraduate Scholarship-Doctoral, Princeton University via a Charlotte Elizabeth Procter Fellowship, and the Miller Institute for Basic Research in Science of UC Berkeley via a Miller Research Fellowship.

## A. Social learning via the replicator dynamics

Social learning can be captured by the replicator dynamics [31–33]. In our case of the 3 × 3 game given by the payoff matrix Eq. 10, the replicator dynamics is

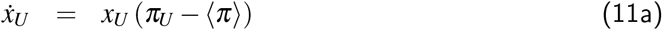

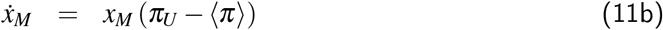

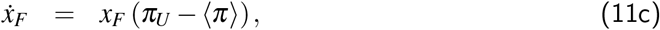

where *x*_*U*_, *x*_*M*_, and *x*_*F*_ are the fractions of individuals using the different strategies (with *x*_*U*_ + *x*_*M*_ + *x*_*F*_ = 1) and dots are derivatives with respect to time. The payoffs are given by

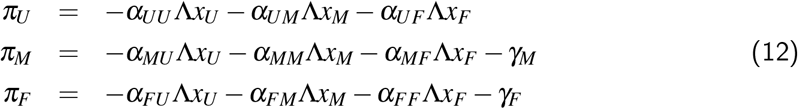

and the average payoff is ⟨ *π* ⟩ = *x*_*U*_ *π*_*U*_ + *x*_*M*_*π*_*M*_ + *x*_*F*_ *π*_*F*_.

For our Fig. 2, we work with Eq. 11a and 11b only and replace *x*_*F*_ by 1 − *x*_*U*_ − *x*_*M*_, which is identically, but numerically more robust. The numerical solutions are computed by Mathematica, the file creating Figure 2 is available at https://zenodo.org/record/7899576.

## B. Game analysis for *α*_*NI*_ − *α*_*II*_ > *α*_*NN*_ − *α*_*IN*_

Here, we analyze the case where the increased infection probability of stopping to follow the intervention in a situation where both have followed is larger than the reduction in infection probability when an individual starts to follow the intervention when both did not, i.e. *α*_*NI*_ − *α*_*II*_ > *α*_*NN*_ − *α*_*IN*_. In this case, the imitation process leads to the following outcomes:

1. When 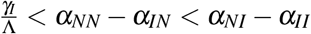, the situation is so risky that adhering to the intervention is optimal for all individuals, resembling a “Harmony game” in game theory.
2. For 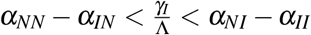, we have a Stag Hunt game: If everybody is adhering to the intervention, one should also follow it. If nobody is adhering to the intervention, one should not start following it. Thus, there are two different stable fixed points of the dynamical (11). In addition, there is an unstable fixed point where the fraction 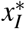 of individuals adhering to the intervention is

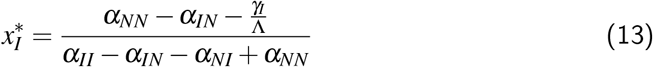 In this situation, universal adherence is the social optimum, but if the initial number of non-adherers is individual too high, the population would converge to an equilibrium where nobody follows the intervention instead.
3. For 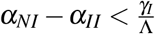, wearing no mask is the dominant strategy:
  a. For 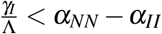, we have a Prisoner’s dilemma – adhering to the intervention is dominated by not adhering, but the social optimum is still that everyone adheres to the intervention.
  b. For 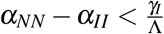, adhering to the intervention is entirely dominated by not adhering – either because the incidence Ξ is low or because the cost *ξ* is low.

## C. Analysing the interaction between two different interventions

Here, we analyse the 3 × 3 game of choosing between masks, (10), in more detail. We start from the situations arising in 3 × 3 games and ask if a third strategy can invade.

### C.1. Invading a situation with no mask usage with *F*

In a situation where one would choose not wearing a mask over an *M* masks, the *F* masks can still be advantageous. This occurs either as a Prisoner’s Dilemma or in the region where using no mask is the equilibrium if only *M* and *U* are considered. Nonetheless, *F* mask use can still be advantageous and spread if *γ*_*F*_ is sufficiently small. This is the case for

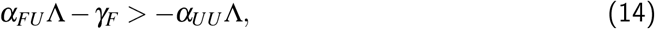

which leads to 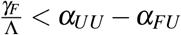.

### C.2. Invasion of a mixture between *U* and *M* by *F*

In a stable mixture between *U* and *M*, the payoff of both strategies is given by

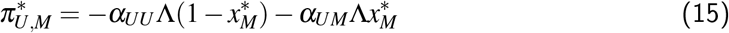

with the fraction of mask users, 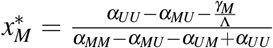, given by (13). In this situation, individuals using *F* masks have a payoff

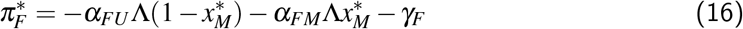

For 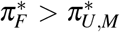, the use of *F* masks would be beneficial in such a coexistence, eventually displacing *M* masks. This is the case for

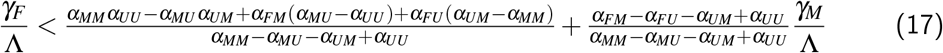

In Fig. 2, this leads to the line that separates the bistability region from other regimes at higher costs *γ*_*F*_.

### C.3. Invasion of a mixture between *U* and *F* by *M*

The *M* masks can also be beneficial in a mixture between *F* and *U*, where the payoff of both strategies is

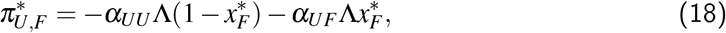

where 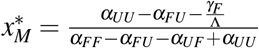. The invasion condition of *M* is

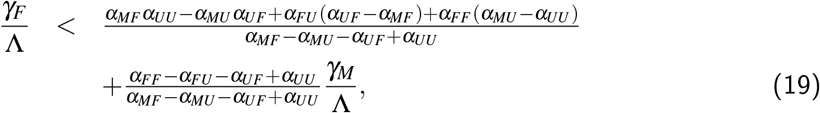

In Fig. 2, this leads to the line that separates the bistability region from the coexistence of *F* and *U* at lower costs *γ*_*F*_.

### C.4. Mixtures between *M* and *F*

For the parameter set given in Table 1, there is no stable coexistence between *M* and *F*. If the risk is sufficiently high, the strategy not to use masks, *U* could always invade when *γ*_*F*_ > *γ*_*M*_. For *γ*_*F*_ < *γ*_*M*_, there is little incentive to use the *M* masks and the whole population would converge on *F* masks if the risk is sufficiently high.

